# Clinical Performance of the STANDARD™ G6PD Test in India

**DOI:** 10.1101/2025.04.03.25324856

**Authors:** Arunansu Talukdar, Santasabuj Das, Sandip Mukherjee, Supriya Sharma, Naseem Ahmed, Maria Kahn, Greg Bizilj, Mikka Nyarko, Benedikt Ley, Ric N Price, Nicole Advani, Emily Gerth-Guyette, Pooja Bansil, Stephanie Zobrist, Gonzalo J Domingo, Abhijit Sharma, Sampa Pal

## Abstract

**Background and objective:** G6PD enzyme deficiency is a genetic condition that can lead to acute hemolysis. This study evaluates the clinical performance of the point-of-care STANDARD™ G6PD Test in Kolkata, India, an area of high incidence of urban *vivax* malaria.

**Methods:** Evaluation of the STANDARD G6PD Test was performed against a reference G6PD assay using UV-spectrophotometry in a febrile population (N=951) recruited from outpatient clinics. Hemoglobin performance was compared to automated hematology analyzer measurements from venous blood. Genotyping was performed on a subset of samples including all deficients/intermediates to identify variants.

**Results:** G6PD deficiency was 2.8%. 20.4% of the study population and had either moderate or severe anemia at enrollment. The test’s sensitivity and specificity were 100% (95% CI: 87.2%– 100%) and 97.8% (96.3%–98.8%), respectively, for G6PD deficient cases. Sensitivity and specificity for females with intermediate G6PD activity were 62.5 % (35.4%–84.8%) and 96.1% (93.3%–98.0%), respectively. Female intermediates with false normal G6PD results had reference G6PD activity greater than 45%. Among malaria-positive individuals (N=272), the test showed 100% sensitivity for both deficient and intermediate G6PD cases. Fifteen participants were identified as Orissa, 1 as Mahidol and 2 as Mediterranean variants. All severe anemia cases (n=24) were accurately categorized as such by STANDARD G6PD test.

**Interpretation and conclusion:** The STANDARD G6PD Test tested at the fever clinic setting was reliable in diagnosing patients at greatest risk of drug induced hemolysis and supports safe treatment of *P. vivax* in this population.

## 1. Introduction

G6PD deficiency is an X-linked disorder that affects about 400 million people worldwide[1]. Males are hemizygous with a genotype of G6PD-deficient or G6PD-normal whereas, females are either homozygous or heterozygous and thus show a wide range of G6PD level from normal to severely deficient.

In tropical areas, G6PD deficiency is estimated at 3–35% [2–6]. According to the 2023 World Malaria Report, global *Plasmodium vivax* (*P. vivax*) malaria cases reached 6.9 million in 2022 and in the region of WHO South-East Asia, 66% of all malaria cases are reported in India [7,8]. The goal towards elimination of malaria in India is particularly challenging in the eastern states, where more than 40% of the country’s malaria occurs [9]. G6PD deficiency was reported in India in the late 90’s with an overall prevalence of 8% and a prevalence of 7.7% in malaria-endemic areas [10–13]. However, recent data on the prevalence of G6PD deficiency is limited.

The 8-aminoquinoline compounds (primaquine and tafenoquine) are the only licensed antimalarial drugs against the dormant liver stages (hypnozoites) of *P.vivax* and *P.ovale* [14,15]. The main concern limiting the use of these drugs is the risk of acute hemolytic anemia in patients with G6PD deficiency [16,17]. The WHO malaria treatment guidelines recommend routine testing for G6PD deficiency prior to primaquine-based radical cure [14,17,18]. However, the inaccessibility of G6PD point of care tests in malaria-burden areas limits the broader clinical use of 8-aminoquinoline radical cure [19,20]. Until recently, especially in low-resource settings, it has not been possible to determine the G6PD status of a patient presenting to a clinic for treatment with a point-of-care test [21,22]. Affordable, appropriate tests for G6PD deficiency has potential to significantly increase access to safe and effective *P. vivax* case management, thereby increasing the effectiveness of malaria elimination [23][24].

The gold standard test for G6PD testing is the complex quantitative spectrophotometry method which requires trained staff and laboratory setting, thus making it challenging for point of care use in malaria-endemic settings [22]. The fluorescent spot test (FST) [25], is a common qualitative test which is low cost and easy to use but also requires laboratory settings, and trained technicians. Moreover, the FST is not suitable for discriminating heterozygous females with intermediate G6PD activity. New lateral flow rapid diagnostic tests (RDTs) for G6PD status also face similar challenges with low accuracy for detecting intermediate G6PD activity [26–29]. Quantitative tests can provide a numerical output of G6PD activity [30] and determine normal, intermediate and/or deficient activity [20–22].

The STANDARD G6PD Test developed by SD Biosensor (South Korea) is a quantitative test for G6PD deficiency that provides a numerical value of G6PD enzyme activity (U/g Hb) and hemoglobin concentration (g/dL) to discriminate G6PD normal from intermediate and deficient as per the thresholds set by SD Biosensor [31,32]. This study determined the diagnostic accuracy of the STANDARD G6PD Test in measuring G6PD level in a febrile cohort in two clinics in Kolkata, India.

## 2. Methods

### 2.1 Ethical considerations

This study was reviewed and approved by the PATH Research Ethics Committee (approval number 1223628), ICMR-NICED Institutional Ethics Committee (approval number A-1-2/2018/IEC), and the Medical College of Kolkata Institutional Ethics Committee (approval number MC/Kol/IEC/Spon/113/06-2018). In addition, the study received approval from the Ministry of Health Screening Committee. Study participants were enrolled after providing written informed consent. For minors below 18 years, consents were obtained from their parents, and written assent was provided by the minor. The study period was 8/29/2019 to 8/8/2022.

### 2.2 Study design

This was a cross-sectional clinical study in Kolkata, India, for diagnostic accuracy. Participants were enrolled at out-patient clinics at the ward-62 Health clinic (HC) and Medical College Hospital Kolkata (MCK). The STANDARD G6PD Test was tested on venous and capillary blood samples followed by validation against the G6PD reference assay using venous samples. Study endpoints included (i) Accuracy of the G6PD activity measurement of the STANDARD G6PD Test on venous and capillary specimens against the G6PD reference spectrophotometric assay, and (ii) accuracy of the hemoglobin concentration measured on the STANDARD G6PD Test on venous and capillary specimens against a hemoglobin reference method.

### 2.3 Study sites

Patients were enrolled at two outpatient clinics at the MCK hospital daily outpatient ward and the ward-62 health fever clinic and outreach center in Kolkata, India. Febrile adults and children 8 years of age or older presenting at recruiting facilities with either an axillary temperature of ≥37.5°C at enrollment or an episode of fever in the last 48 hours were eligible for participation.

### 2.4 Workflow

At the point-of-care, demographic information was collected using a questionnaire. Fresh capillary blood samples were collected by fingerstick and tested with the STANDARD G6PD Test, the HemoCue 201, and a standard malaria RDT. Next, 3mL of venous blood was drawn in K_2_-EDTA tubes and transported in cold chain to the National Institute of Cholera and Enteric Diseases (ICMR-NICED) laboratory in Kolkata, where the G6PD reference assay, the STANDARD G6PD Test and HemoCue tests were performed in the laboratory using the venous specimens (Table 1). Technicians using the investigational test were blinded to the reference assay test results, and vice versa. Remaining venous blood K_2_-EDTA specimens were frozen at – 80C at NICED. For G6PD mutation analysis, a subset of frozen specimens was sent to the National Institute of Malaria Research (ICMR-NIMR) in New Delhi, India.

**Table 1.**
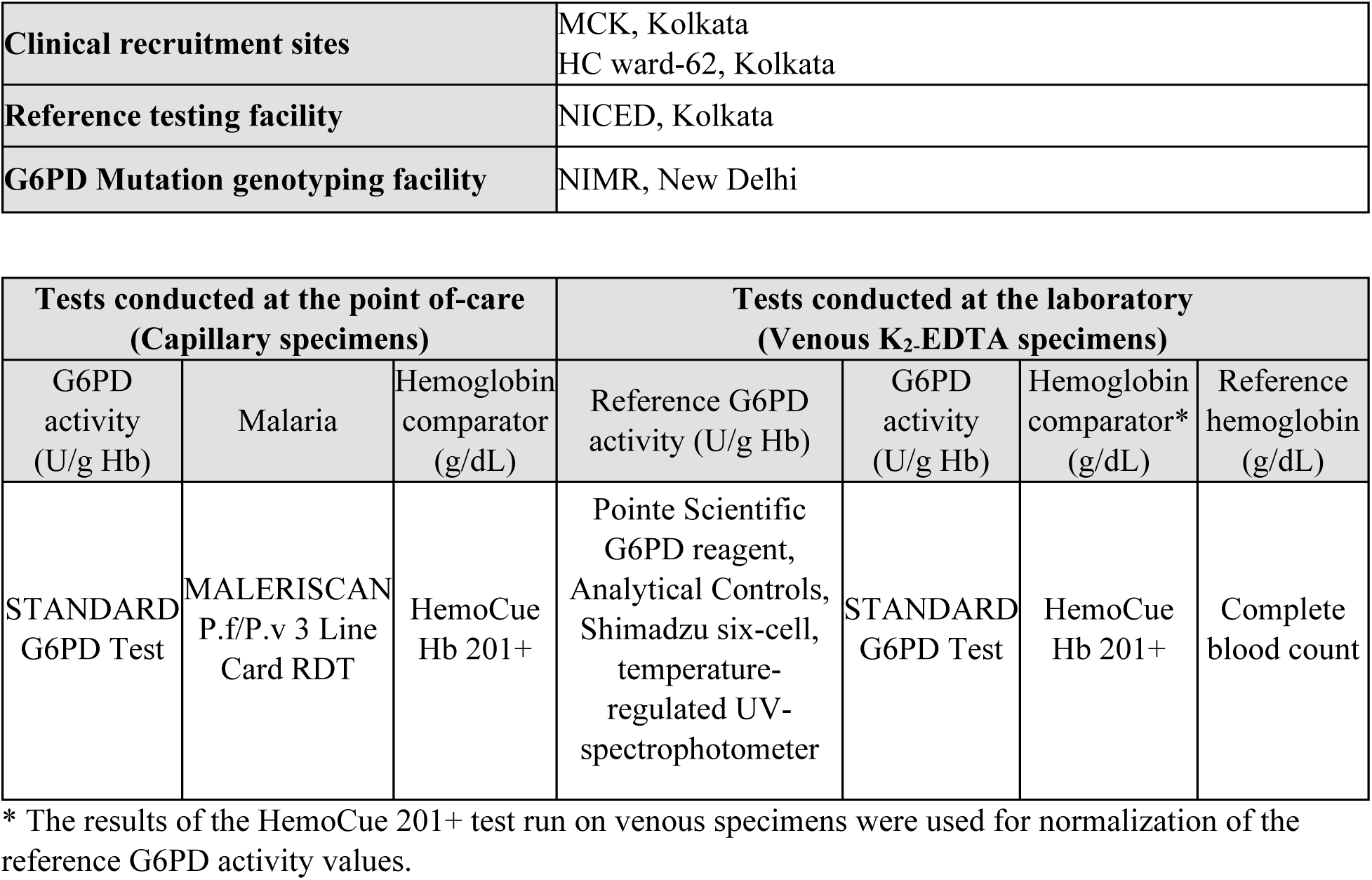
Summary of study tests.

|  |  |
| --- | --- |
| Clinical recruitment sites | MCK, Kolkata<br>HC ward-62, Kolkata |
| Reference testing facility | NICED, Kolkata |
| G6PD Mutation genotyping facility | NIMR, New Delhi |

| Tests conducted at the point-of-care<br>(Capillary specimens) |  |  | Tests conducted at the laboratory<br>(Venous K<sub>2</sub>EDTA specimens) |  |  |  |
| --- | --- | --- | --- | --- | --- | --- |
| G6PD activity<br>(U/g Hb) | Malaria | Hemoglobin<br>comparator<br>(g/dL) | Reference G6PD<br>activity (U/g Hb) | G6PD activity<br>(U/g Hb) | Hemoglobin<br>comparator*<br>(g/dL) | Reference<br>hemoglobin<br>(g/dL) |
| STANDARD<br>G6PD Test | MALERISCAN<br>P.f/P.v 3 Line<br>Card RDT | HemoCue<br>Hb 201+ | Pointe Scientific<br>G6PD reagent,<br>Analytical Controls,<br>Shimadzu six-cell,<br>temperature-<br>regulated UV-spectrophotometer | STANDARD<br>G6PD Test | HemoCue<br>Hb 201+ | Complete<br>blood count |

\* The results of the HemoCue 201+ test run on venous specimens were used for normalization of the reference G6PD activity values.

### 2.5 Point-of-care testing (capillary specimens)

At the clinics, the following tests were conducted on non-anticoagulated capillary specimens (Table 1):

#### Malaria test

MALERISCAN RDT MALARIA P.f/P.v 3 Line Antigen from Bhat Biotech, India was used for routine qualitative detection of HRP II (Histidine Rich Protein 2) and pLDH (*Plasmodium* lactate dehydrogenase) antigens in capillary blood.

#### Comparator hemoglobin test

The HemoCue^®^ Hb 201+ System (HemoCue AB, Sweden) was used to measure the concentration of hemoglobin in g/dL as a comparator.

#### STANDARD G6PD Test

The STANDARD G6PD Test measured the G6PD activity value by normalized hemoglobin concentration and provided enzyme activity in units per gram hemoglobin (U/g Hb) and a value for total hemoglobin (T-Hb) concentration in g/dL. The STANDARD G6PD Test includes test kits and an analyzer. The test kit consists of test devices, a code chip specific to each lot, extraction buffer tubes, Ezi+ tube for blood collection, and Instructions for Use. Thresholds for G6PD activity have been established by the manufacturer for the classification of G6PD status based on the test’s numeric results outputs: >6.0 U/g Hb for G6PD-normal females (>70% G6PD activity), >4.0 to 6.0 U/g Hb for G6PD-intermediate females (>30% to ≤70% G6PD activity), >4.0 U/g Hb for G6PD-normal males (>30% G6PD activity), and ≤4.0 U/g Hb for G6PD-deficient males and females (≤30% G6PD activity) [33].

To perform the test, a test device is first placed into the analyzer. Next using the Ezi+ tube, 10µL of blood specimen is transferred to the extraction buffer tube and mixed. The blood-buffer solution is then collected using a new Ezi tube+ and applied to the test strip. Test runs for 2 minutes. The analyzer shows the result of G6PD activity in U/g Hb and hemoglobin concentration in g/dL. For quality assurance, high and low G6PD unitized controls are available from SD Biosensor. The control reagents (02G6C10, SD Biosensor, Republic of Korea) are lyophilized pellets, with a workflow similar to sample testing, using the device in control mode.

### 2.6 Laboratory testing (venous blood)

At the ICMR-NICED laboratory, venous K_2-_EDTA blood specimens were aliquoted and used to conduct further G6PD and hemoglobin testing as described below (Table 1).

#### STANDARD G6PD Test

The test was used as described above following the user instructions manual.

#### Reference G6PD activity assay

G6PD activity was measured using the quantitative G6PD kit from Pointe Scientific (MedTest, Canton, MI, USA, catalog number G7583) on a six-cell, temperature-controlled UV spectrophotometer (Shimadzu UV-1800, Japan). The Pointe Scientific assay was performed at 37°C and G6PD activity values were normalized using the venous HemoCue hemoglobin concentration and expressed in U/g Hb. Each day, normal, intermediate, and deficient controls (Analytical Control Systems Inc. Indiana, USA, catalog numbers HC-108, HC-108IN, and HC-108DE, respectively) were run for the reference assay.

#### Comparator hemoglobin test

The HemoCue^®^ Hb 201+ test was performed on venous K_2-_EDTA blood as a comparator. The results from the test were computed to normalize the G6PD reference assay result.

#### Reference hemoglobin assay

Hemoglobin testing with a complete blood count was also available for a subset of 481 specimens using an automated hematology analyzer (DxH 900 Beckman Coulter) and was considered as the reference assay for hemoglobin.

#### Sanger sequencing for G6PD mutations

G6PD mutation analysis was performed at NIMR for a subset of 107 specimens. G6PD-deficient and intermediate specimens as well as discordant samples were selected for sequencing, along with 21 randomly selected G6PD-normal specimens according to the reference spectrophotometry assay for a total of 107 specimens. Isolation of Genomic DNA from thawed frozen venous samples was performed using QIAamp Mini Kit (catalog number 51104, Germany). Isolated DNA was stored at −20°C until used for the genotyping analysis. Amplification of the G6PD exons and DNA sequencing was performed as previously reported [34,35]. Specific primers were used for the amplification of the most frequent G6PD mutant type found in India (i.e., G6PD Mediterranean [C563T], G6PD Orissa [C131G], G6PD Kerala-Kalyan [G949A], and G6PD Mahidol [G487A]). All PCR products were subjected to DNA sequencing (Eurofin, India). DNA sequencing data was analyzed using MEGA10 software. To identify the mutations, the sequences were compared to the G6PD gene ensemble (Accession No. ENSG00000160211).

### 2.7 Operating temperature

Thermometers and hygrometers were employed at both the point of care and in the laboratory to collect temperature and humidity information during testing.

### 2.8 Data management

Results were collected on case report forms and data was recorded into the REDCap^®^ (Research Electronic Data Capture) system [36], which is a secure, web-based application that includes built-in validation rules to minimize errors. Data was monitored regularly against the source documents for accuracy.

### 2.9 Statistical methods

In order to normalize the reference assays’ absolute G6PD values, the adjusted male median (AMM) was established from 36 normal males that are randomly selected [27][37]. This subset was excluded from further analysis of performance. Using the AMM (100% activity), G6PD normal individuals were defined as males with >30% and females with >70% activity, G6PD deficient participants as males and females with ≤30% activity, and intermediate females as those with >30-70% activity levels. For the STANDARD G6PD Test, the manufacturer’s recommended thresholds of 6.0 U/g Hb and 4.0 U/g Hb were applied to the test results to group participants as G6PD normal, intermediate, and deficient based on sex, as described above. The sensitivity and specificity of the STANDARD G6PD Test was calculated against the reference assay according to standard methods and reported with 95% confidence intervals [19]. Sensitivity and specificity of the STANDARD G6PD Test are presented separately for capillary and venous specimens against the reference assay result from venous specimens.

Receiver operating characteristic (ROC) curves differentiated G6PD females and deficient males at the 70% and 30% thresholds. Linear regression was performed to measure the correlation of G6PD activity between the STANDARD G6PD Test and the reference assay. Additionally, the agreement between the STANDARD G6PD Test and the reference assay was determined for the classification of G6PD status.

Similarly, correlation was determined for the hemoglobin concentration between capillary and venous STANDARD G6PD Test results and reference hemoglobin values from the venous CBC results. Bland-Altman plots are also presented to describe the mean difference between experimental and reference assay in g/dL for the hemoglobin measurements. Bias in hemoglobin measurement was also calculated as the mean difference between the hemoglobin concentration as determined on the STANDARD G6PD Test, minus the mean on the reference hemoglobin assay. Percent agreement between the two methods was also assessed for the classification of anemia status using WHO definitions [31,38]. No anemia and mild anemia were categorized together given the clinical importance of severe and moderate anemia. Lastly, given the clinical significance of severe anemia—specifically for *vivax* patients receiving 8-aminoquinolines—the diagnostic performance of the STANDARD G6PD Test was also calculated for the identification of severely anemic cases for sensitivity, specificity, negative predictive value (NPV) and positive predictive value (PPV). All analyses are also presented for the HemoCue results on capillary and venous specimens against the reference assay, as a comparator.

Stata 13.0 and 15.0 (StataCorp, College Station, TX, USA) were used for statistical analysis.

The data generated from this India study contributed to the pooled analysis[31]. However, the pooled analysis does not include an in-depth analysis of the India specific study data including the analysis of the underlying G6PD genotypes.

## 3. Results

### 3.1 Study Population

951 participants were enrolled and included in the analytical population. The mean age of the study population was 36.8 years, ranging from 8 to 81 years. A total of 63.7% (n=606) of participants were males and 36.3% (n=345) were females. 20.4% of the population had either moderate or severe anemia, and 28.6% tested positive for malaria at enrollment. Nearly all (94.4%) of the *P. vivax-*positive participants were recruited from the health clinic ward-62. No significant differences in median or mean hemoglobin concentrations were observed between *P. vivax* malaria positive/negative participants, or G6PD normal/deficient participants.

### 3.2 Distribution of G6PD activity

The study’s AMM was calculated at 8.56 U/g Hb, which was used to define 100% activity. Accordingly, the reference assay 70% and 30% activity thresholds corresponded to 5.99 U/g Hb and 2.57 U/g Hb, respectively. These thresholds give a total of 27 G6PD deficient males and females and 17 intermediate females in the study population. The distribution of G6PD activity by sex in percent activity is shown in Figure 1.

**Figure 1.**
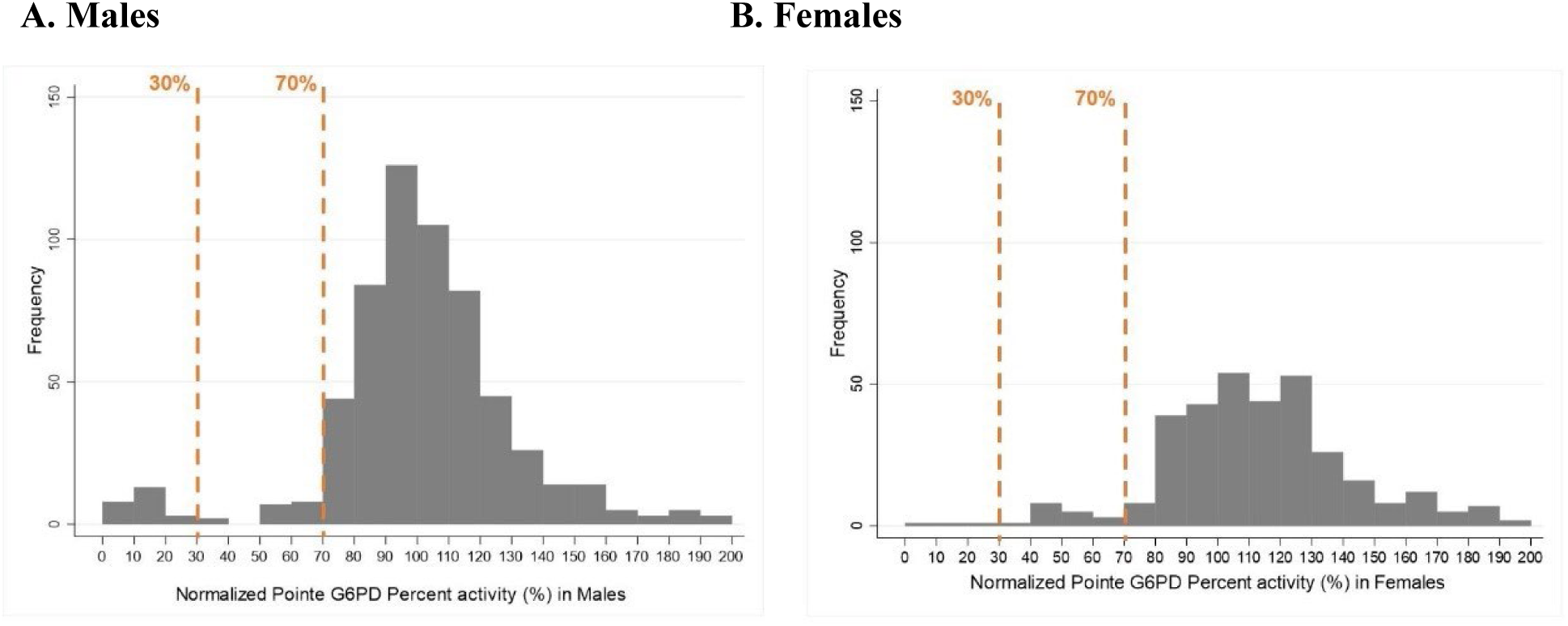
Male and female G6PD activity distribution in percent activity as measured by spectrophotometry.

### 3.3 Operation of the STANDARD G6PD Test

Out of 951 participants, STANDARD G6PD Test results were excluded for 57 capillary specimens and 66 venous specimens due to QC being performed with expired control reagents. G6PD activity distributions on the STANDARD G6PD Test are shown in Supplementary Figure 1 STANDARD G6PD Test was conducted under temperatures ranging from 17.1°C to 38.9°C (Supplementary Table 1), which is representative of expected conditions and within the operating temperatures of the STANDARD G6PD Test.

### 3.4 ROC curves for the STANDARD G6PD Test for G6PD

Figure 2 shows the results of the ROC curve analysis for capillary specimens. An AUC of 0.996 was observed for G6PD deficient males and females (A), and 0.884 for G6PD intermediate females (B). Similar results were observed with venous specimens (Supplementary Figure 2).

**Figure 2.**
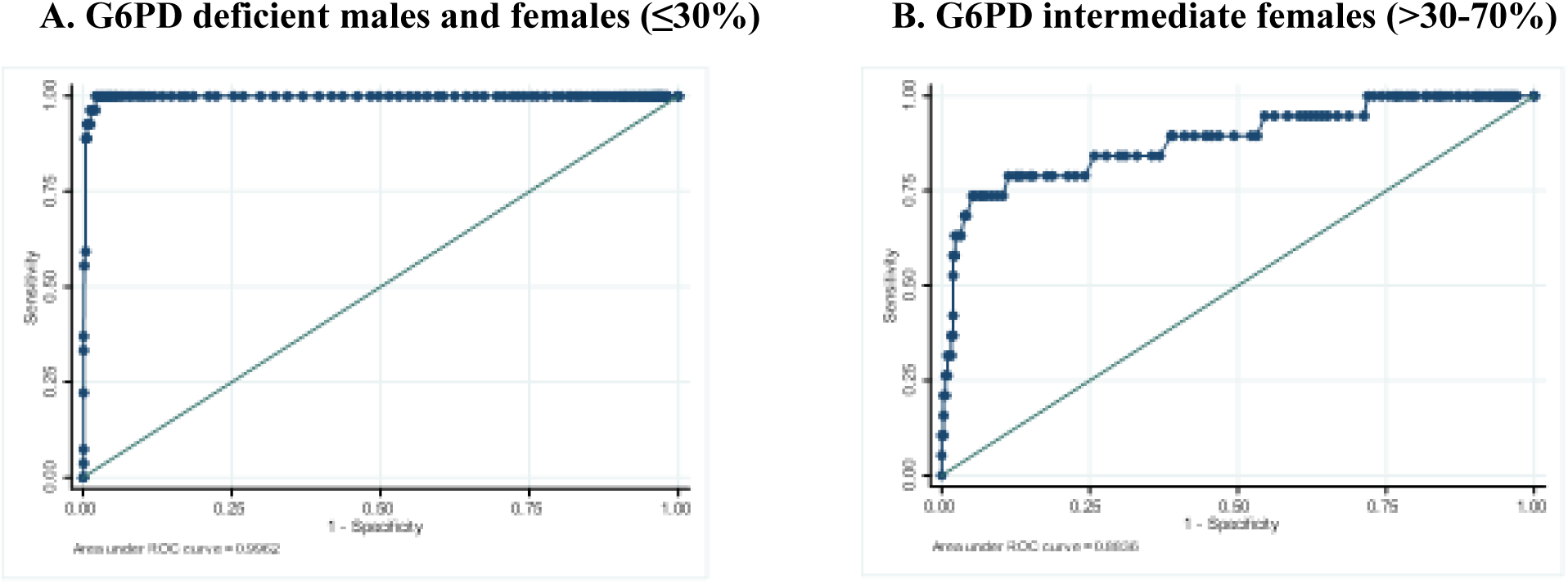
ROC curves for the STANDARD G6PD measurement on capillary specimens and the test’s ability to discriminate (A) G6PD normal males and females from deficient male and female, and from intermediate females; (B) Intermediate female G6PD activity (> 30%–70%) from normal females.

### 3.5 STANDARD G6PD Test Performance

The performance analysis showed 100% sensitivity for deficient males and females with G6PD activity ≤ 30% G6PD across capillary and venous specimens, and a specificity of >95%. The test also demonstrated 100% sensitivity in all malaria positive cases for deficient individuals and intermediate females using both venous and capillary specimens. Among all female G6PD intermediates, the test demonstrated > 62% sensitivity with a specificity >94% across both specimen types (Table 2). two-by-two tables for all diagnostic performance comparisons are provided in the supplementary materials (Supplementary Tables 2 to 9).

**Table 2.** Performance of the STANDARD G6PD Test.

|  | Venous specimens | Capillary specimens |
| --- | --- | --- |
| **Overall study population** |  |  |
| $\leq 30\%$ G6PD-deficient males and females, total study number | 859 | 888 |
| Sensitivity (95% CI) | 100.0 (86.8–100.0) | 100.0 (87.2–100.0) |
| Specificity (95% CI) | 97.6 (96.3–98.5) | 97.6 (96.3–98.5) |
| $> 30\text{--}70\%$ intermediate females, total study number | 321 | 326 |
| Sensitivity (95% CI) | 64.7 (38.3–85.8) | 62.5 (35.4–84.8) |
| Specificity (95% CI) | 94.1 (90.8–96.5) | 96.1 (93.3–98.0) |
| **Malaria positive cases** |  |  |
| $\leq 30\%$ G6PD-deficient males and females, total study number | 221 | 232 |
| Sensitivity (95% CI) | 100.0 (29.2–100.0) | 100.0 (39.8–100.0) |
| Specificity (95% CI) | 98.2 (95.4–99.5) | 96.9 (93.8–98.8) |
| > 30%–70% intermediate females, total study number | 43 | 42 |
| Sensitivity (95% CI) | 100.0 (2.5–100.0) | 100.0 (2.5–100.0) |
| Specificity (95% CI) | 92.9 (80.5–98.5) | 95.1 (83.5–99.4) |

Six females with intermediate G6PD activity showed false normal on the STANDARD G6PD Test. results, particularly from specimens with G6PD activity levels close to the threshold of upper intermediate (70%). Table 3 shows the predictive power, sensitivity, and specificity for deficient and intermediate females with G6PD activity levels <45%, <50%, <60%, <65%, and <70%. Only two females with false normal STANDARD G6PD Test results above 6.0 U/g Hb had G6PD activity levels <50% on the reference assay (and all were greater than 45%).

**Table 3.**
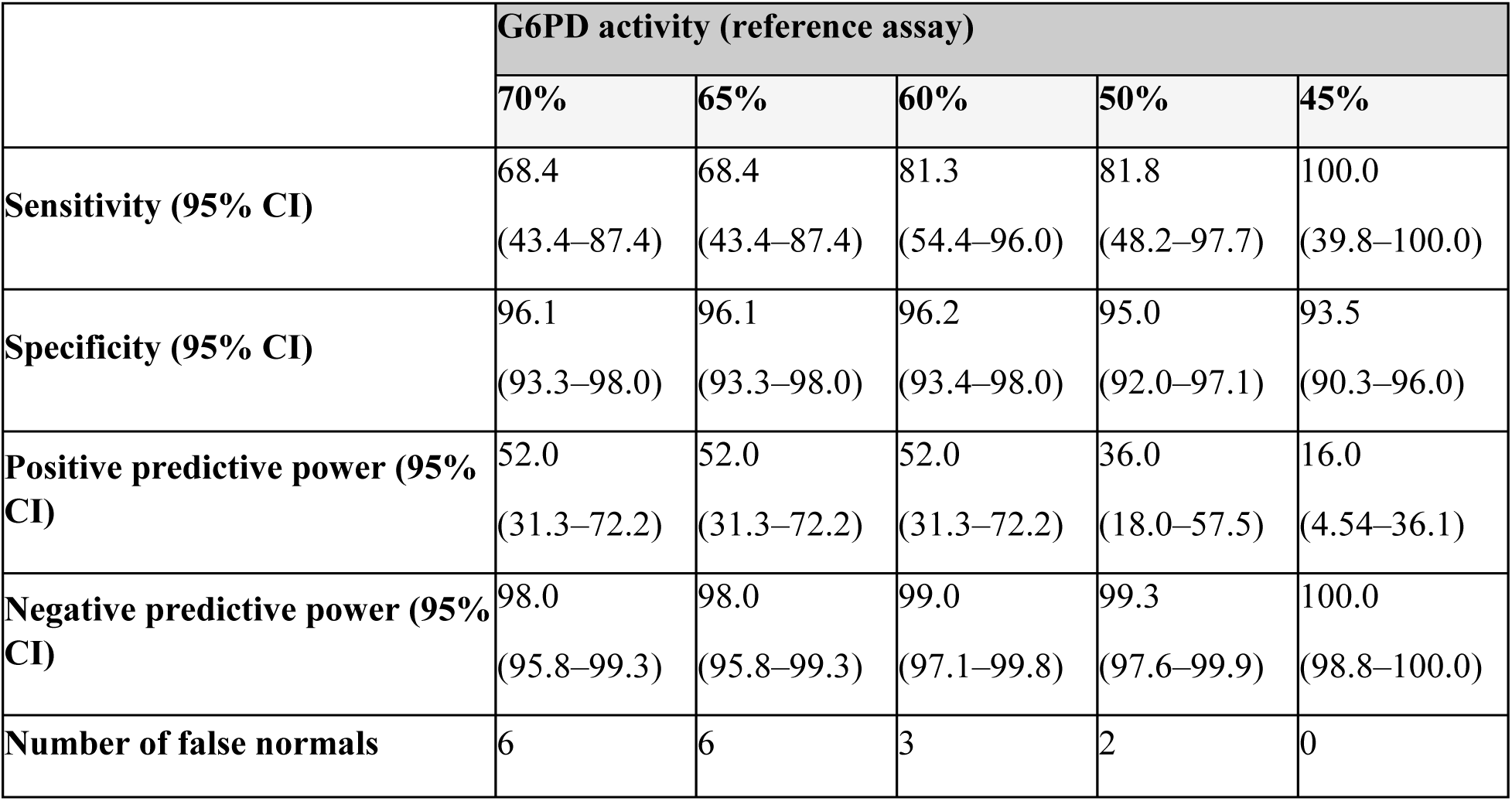
STANDARD G6PD Test performance for capillary specimens, all females (N=326) on the 6.0 U/g Hb threshold.

The overall percent agreement of the STANDARD G6PD Test against the reference assay’s result classifications for G6PD normal, intermediate, and deficient individuals was calculated for each specimen type. The percent agreement of the G6PD activity reference assay and the STANDARD G6PD Test categorized results on venous and capillary specimens was 95.2% [95% CI: 93.6–96.6] and 96.2% [95% CI: 94.7–97.3], respectively (Supplementary Tables 10 and 11).

### 3.6 Correlation of G6PD activity between the STANDARD G6PD Test and reference assay

Correlation of the G6PD activity between STANDARD G6PD Test and reference assay results showed R-squared values of 0.66 for capillary and 0.57 for venous specimens by regression analysis (Figure 3). Higher imprecision was noted for G6PD values in the high or normal G6PD activity range. The same correlations in the sub-population of only the malaria-infected participants showed lower correlation values of 0.59 for capillary and 0.41 for venous specimens (Supplementary Figure 3).

**Figure 3.**
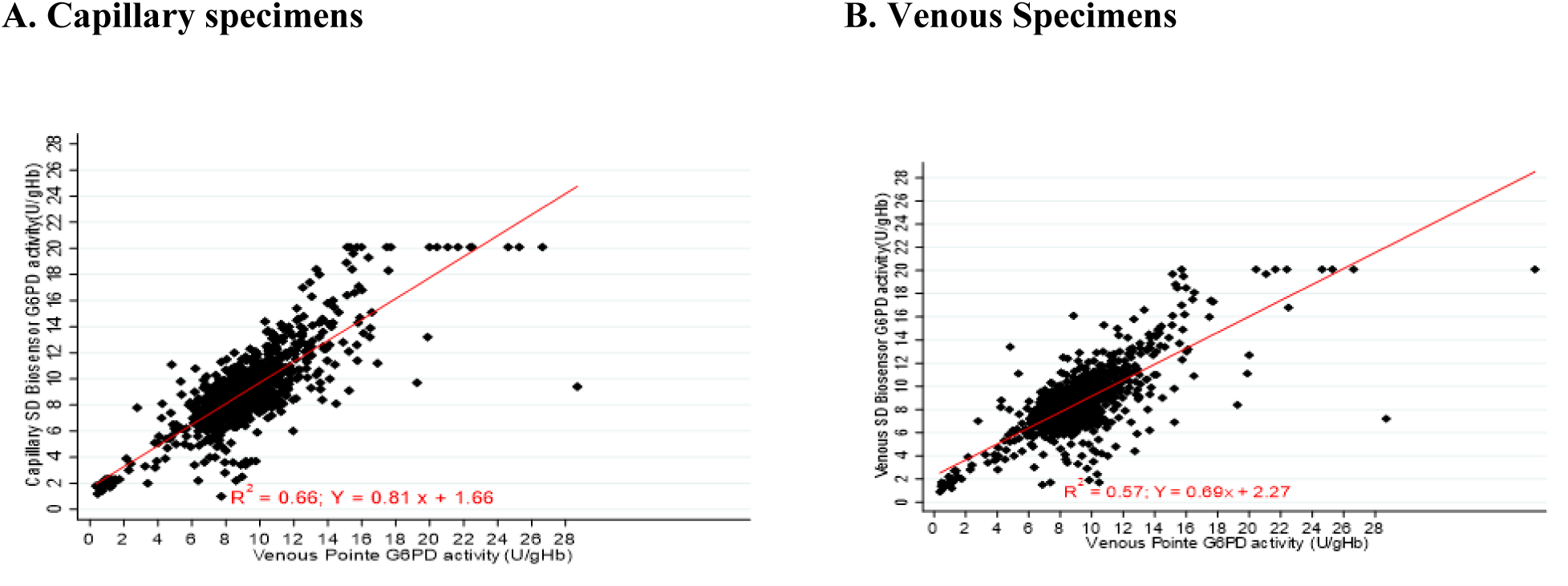
Correlation of the G6PD activity on STANDARD G6PD Test, capillary (A) and venous blood (B) and reference assay, venous blood.

**Figure 4.**
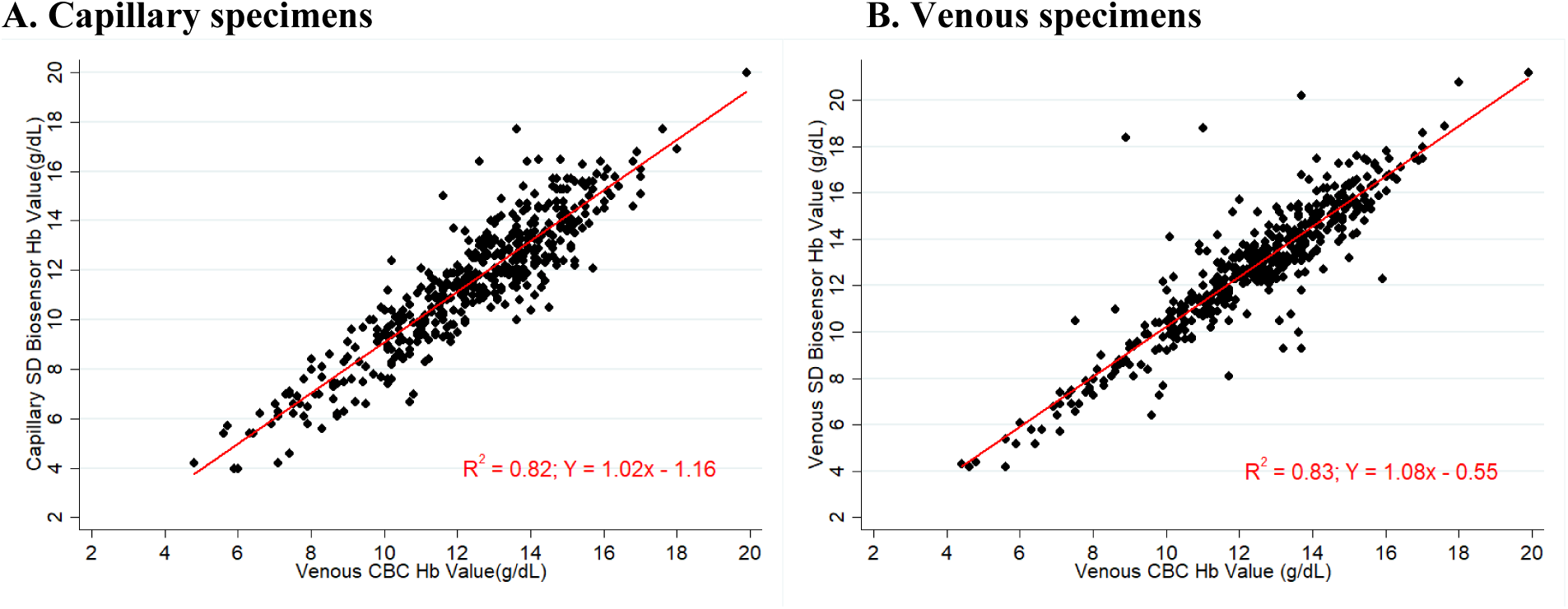
Linear regression analysis of the STANDARD G6PD Test hemoglobin concentration result for capillary (A) and venous blood (B) against the reference CBC result from venous blood.

### 3.7 STANDARD G6PD Test performance for hemoglobin

The hemoglobin results of capillary and venous specimens on the STANDARD G6PD Test were compared to the reference CBC result on venous specimens through linear regression and Bland Altman plots (Supplementary Figure 4). The R-square values for capillary and venous were 0.82 and 0.83, respectively. Bias (difference in means) between the reference CBC hemoglobin and STANDARD G6PD Test values were −0.85 g/dL (standard deviation [SD] 1.10) on capillary specimens and 0.43 g/dL (SD 1.21) on venous specimens (Supplementary Table 12). In contrast, the R-square values for the HemoCue were slightly higher at 0.85 and 0.93 for capillary and venous specimens, respectively (Supplementary Figure 5). However, mean differences were similar at −0.10 (SD 1.04) for capillary and 0.19 (SD 0.70) for venous (Supplementary Table 12).

Percent agreement was compared between the reference CBC and STANDARD G6PD Test’s classification of anemia status on both venous and capillary specimens based on the device’s hemoglobin concentration result (Table 4). Percent agreement between the reference CBC and STANDARD G6PD Test’s anemia status on capillary specimens was 80.5% 95% [CI: 76.6– 84.0]; on venous specimens, the percent agreement was 90.6% [95% CI: 87.5–93.1]. No cases of severe anemia were misclassified by the STANDARD G6PD Test as no or mild anemia on either capillary or venous specimens. The HemoCue showed similar patterns of misclassification, with an overall percent agreement of 90.0% for capillary specimens and 96.9% for venous specimens (Supplementary Table 13). In terms of diagnostic performance for the identification of severe anemia, the STANDARD G6PD Test showed slightly higher sensitivity than the HemoCue for the identification of severely anemic cases on both specimen types, though the PPV is lower (Supplementary Table 14).

**Table 4.** Agreement between the STANDARD G6PD Test and reference CBC’s hemoglobin concentration (in g/dL) -based anemia classifications as per WHO definitions on (A) capillary and (B) venous specimens.

A. Capillary
|  |  | CBC (venous) |  |  |  |
| --- | --- | --- | --- | --- | --- |
|  |  | Non-/mild anemia | Moderate anemia | Severe anemia | Total |
| STANDARD G6PD Test (capillary) | No/mild anemia | 290 | 3 | 0 | 293 |
|  | Moderate anemia | 64 | 60 | 0 | 124 |
|  | Severe anemia | 0 | 24 | 25 | 49 |
| Total |  | 354 | 87 | 25 | 466 |

B. Venous
|  |  | CBC (venous) |  |  |  |
| --- | --- | --- | --- | --- | --- |
|  |  | Non-/mild anemia | Moderate anemia | Severe anemia | Total |
| **STANDARD G6PD**<br>**Test (venous)** | **No/mild anemia** | 327 | 18 | 0 | 345 |
|  | **Moderate anemia** | 18 | 60 | 1 | 79 |
|  | **Severe anemia** | 0 | 6 | 25 | 31 |
| **Total** |  | 345 | 84 | 26 | 455 |

673 Percent agreement: 90.6% (95% CI: 87.5% – 93.1%)

Separately, associations between anemia and G6PD deficiency as well as malaria status were investigated, for the full study population. and none observed (Supplementary Tables 15 and 16).

### 3.8 Genotyping of STANDARD G6PD Test results

Sanger sequencing of genomic DNA and molecular analysis were completed for the G6PD Orissa, Kerala Kalyan, Mediterranean and Mahidol variants on 107 specimens. In total, 18 study participants were identified as carrying one of three common mutations (Supplementary Tables 17): 15 participants with the Orissa variant (131C > G), 2 with the G6PD Mediterranean variant (563C > T), and 1 with G6PD Mahidol (487G > A). The Kerala Kalyan mutation was not detected in any specimens. Of the 27 participants confirmed as G6PD deficient by all three enzyme activity measurements (the reference assay and both capillary and venous STANDARD G6PD Test results), 10 were confirmed G6PD deficient by genotyping: 6 males and 1 homozygous female with the Orissa variant, 2 males with the Mediterranean variant, and 1 male with the Mahidol variant (Supplementary Table 18). No mutations were detected in the 46 specimens that were normal by all tests.

## 4. Discussion

Prevalence of G6PD deficiency is very heterogenous in India, with some populations (and especially tribal populations) experiencing a higher prevalence than others [11,39,40]. This presents a barrier for the *P. vivax* radical cure and to safe access of treatment options with primaquine and tafenoquine [41,42]. This study demonstrates the diagnostic accuracy of the STANDARD G6PD Test when used in malaria-endemic urban setting in eastern India by health care staff. Results demonstrated that the STANDARD G6PD Test accurately identified all G6PD deficient cases with ≤30% G6PD by the reference assay, when conducted both at the outpatient clinics on finger stick specimens as well as when performed with venous K_2_-EDTA specimens in a laboratory. The test’s specificity for G6PD deficient cases was greater than 97% both on venous and capillary specimens. For G6PD intermediate females, the STANDARD G6PD Test also showed > 62% sensitivity across all intermediate females, as well as 100% sensitivity in *P.vivax* cases with a specificity of >92% across venous and capillary specimens. Only two intermediate females with reference G6PD activities < 55% (49% and 50%) were misclassified as normal. Among confirmed *P. vivax* cases, there were no false normal G6PD cases, neither true deficient nor intermediates on the STANDARD G6PD Test, albeit there were low numbers of G6PD deficient and intermediate cases in this sample set.

A subset of the specimens (n=107) were genotyped for the Orissa, Kerala Kalyan, Mediterranean, and Mahidol G6PD variants. Among the 18 study participants identified with a deficient G6PD variant, the predominant observed variant was Orissa (n=15), followed by the Mediterranean (n=2), and Mahidol (n=1). While no cases identified as G6PD normal by all three enzyme activity measurements displayed any of these four variants, genotyping was only able to confirm 10/27 G6PD deficient cases identified by the reference assay. This reflects the limitations of G6PD genotyping without conducting full gene sequencing in India given the complexity of G6PD deficiency in this context. A study completed in Bangladesh in which a subset of samples were genotyped for the G6PD variants (Mahidol, Viangchan, Mediterranean, Orissa, and Kalyan-Kerala) was also unable to resolve all G6PD deficient and intermediate cases through genotyping [43].

Overall, the performance of the test is similar to previously published results albeit with the sensitivity for intermediate females being lower [30,31,33,44]. A recent multi-country pooled performance analysis also confirms that the STANDARD G6PD Test supports safe access to primaquine for individuals with G6PD deficiency [45]. Some of the females classified as intermediate on the reference assay may have resulted from compromised G6PD enzyme activity as a consequence of specimen shipment from the two clinics to the laboratory where reference testing was conducted. Operationally, despite the high specificity, due to low prevalence of G6PD deficiency in Kolkata, there is a high likelihood that a G6PD deficient case by the STANDARD G6PD Test may actually be normal or intermediate (low positive predictive power) in this study. Such cases still represent only 2% (21/888) of the study population that would not have access to 14-day primaquine based on the STANDARD G6PD Test point-of-care results [45]. The STANDARD G6PD Test has shown good user acceptance across multiple settings, and several studies have now demonstrated operational use of the test for management of malaria cases [45–48]. The specificity of the test for intermediate cases in particular seems to decrease under real-world implementation conditions, resulting in a higher proportion of false deficient and intermediate cases. Further operational research should be conducted to understand and identify strategies to minimize or mitigate this observation. Additionally, the high negative predictive power (100% for G6PD normal cases), may present an opportunity in some contexts to significantly reduce re-testing for the majority which are G6PD normal. A recent study also indicates the utility of the test to screen newborns for G6PD deficiency (albeit with higher thresholds) [49], which is also of relevance in some Indian populations with higher G6PD deficiency rates [50,51].

In this study population, a high proportion of participants (20.4%) presented with either moderate or severe anemia at enrollment, based on the results of the HemoCue. Anemia is commonly associated with malaria infection[51], and in particular, *P. vivax* infection is considered a risk factor for severe anemia and poor nutrition outcomes [52–54]. When compared against the reference CBC assay, the HemoCue showed higher performance for the measurement of hemoglobin than the STANDARD G6PD Test on both venous and capillary specimens. However, bias between both index and reference tests was similar (Supplementary Table 13), and both tests showed similar patterns of misclassification, with no cases of severe anemia being misclassified by either test as no or mild anemia on any specimen type (Table 4, Supplementary Table 14). Indeed, when considering diagnostic performance of both tests for the identification of severely anemic cases, the STANDARD G6PD Test showed slightly higher sensitivity than the HemoCue for the identification of severely anemic cases on both specimen types, though with a lower PPV (Supplementary Table 15). These results are consistent with the findings from other performance evaluations of this test [45] and suggest that the STANDARD G6PD Test can reliably identify cases of severe anemia.

In brief, this study demonstrated the use and performance of the STANDARD G6PD Test at two outpatient urban point-of-care clinics in Kolkata, India, where malaria is treated, and anemia is prevalent. From a performance perspective, the test eliminated the risk of severe G6PD deficient cases being exposed to drugs for which G6PD deficiency is a contra-indication, as well as significantly reducing the risk for females with low intermediate G6PD deficiency. As such, the STANDARD G6PD Test can improve safe access to 8-aminoquinolines for the *P. vivax* radical cure in Indian populations with high prevalence of G6PD deficiency.

## Data Availability

All data produced in the present study are available upon reasonable request to the authors and also contained in the manuscript.

## Supplementary Materials

Supplementary Table 1: Operating temperatures of the STANDARD G6PD Test by site and specimen (degrees Celsius). Supplementary Table 2. **S**ensitivity and specificity: 2 x 2 table of the STANDARD G6PD Test against the G6PD reference assay for G6PD deficient venous specimens. Supplementary Table 3. **S**ensitivity and specificity: 2 x 2 table of the STANDARD G6PD Test against the reference G6PD assay for G6PD intermediate (> 30%–70%), female venous specimens. Supplementary Table 4. **S**ensitivity and specificity: 2 x 2 table of the STANDARD G6PD Test against the G6PD reference assay for G6PD deficient capillary specimens. Supplementary Table 5. Sensitivity and specificity: 2 x 2 table of the STANDARD G6PD Test against the G6PD reference assay for G6PD intermediate (> 30%– 70%), female capillary specimens. Supplementary Table 6. Sensitivity and specificity: 2 x 2 table of the STANDARD G6PD Test against the G6PD reference assay for G6PD deficient venous specimens (malaria positives only). Supplementary Table 7. Sensitivity and specificity: 2 x 2 table of the STANDARD G6PD Test against the G6PD reference assay for G6PD intermediate (> 30%–70%), female venous specimens (malaria positives only). Supplementary Table 8. Sensitivity and specificity: 2 x 2 table of the STANDARD G6PD Test against the G6PD reference assay for G6PD deficient capillary specimens (malaria positives only). Supplementary Table 9. Sensitivity and specificity: 2 x 2 table of the STANDARD G6PD Test against the G6PD reference assay for G6PD intermediate (> 30%–70%), female capillary specimens (malaria positives only). Supplementary Table 10. G6PD status classification: agreement between the STANDARD G6PD Test and the G6PD reference assay using venous specimens. Supplementary Table 11. G6PD status classification: agreement between the STANDARD G6PD Test and the G6PD reference assay using capillary specimens. Supplementary Table 12. Mean difference and standard deviation by specimen type, for the STANDARD G6PD Test and the HemoCue hemoglobin measurements, compared to the reference complete blood count (CBC) assay. Supplementary Table 13. Anemia status classification: agreement between the HemoCue and reference CBC’s hemoglobin concentration (in g/dL) -based anemia classifications as per WHO definitions on (A) capillary and (B) venous specimens. Supplementary Table 14. Diagnostic performance of the STANDARD G6PD Test and the HemoCue comparator for the identification of severe anemia on (A) capillary and (B) venous blood against the reference complete blood count (CBC) assay on venous blood. Supplementary Table 15. Anemia status‡ of the study population, by malaria status. Supplementary Table 16. Anemia status of the study population, by G6PD status. Supplementary Table 17. Primer sequence for primers used to amplify G6PD exons. Supplementary Table 18. Study participants with confirmed G6PD mutations as determined by genotyping. Supplementary Figure 1. G6PD activity distributions of study participants on the STANDARD G6PD Test for (A) males-capillary, (B) males-venous, (C) females-capillary, and (D) females-venous. Supplementary Figure 2. ROC curves for the STANDARD G6PD measurement on venous specimens and the test’s ability to differentiate (A) G6PD normal and deficient males, females from female intermediate; (B) female intermediate with G6PD > 30%–70% from normal females. Supplementary Figure 3. Correlation of STANDARD G6PD Test by linear regression analysis: G6PD activity on capillary (A) and venous blood (B) versus the Pointe Scientific reference assay on venous blood for malaria positive cases. Supplementary Figure 4. Correlation of STANDARD G6PD Test hemoglobin results by linear regression analysis and Bland Altmans on capillary (A) and venous blood (B) against the reference complete blood count (CBC) on venous specimens as the reference assay. Supplementary Figure 5. Correlation of the HemoCue hemoglobin results by linear regression analysis and Bland Altmans on capillary (A) and venous blood (B) against the reference complete blood count (CBC) on venous specimens as the reference assay.

## Informed Consent Statement

Patients gave approval and signed informed-consent forms for the participation.

## Acknowledgments

We thank all the health care workers and participants in the study.

