## Supplemental Materials for "Clinical Performance of the STANDARD™ G6PD Test in India"

### Supplementary Materials

**Supplementary Table 1**. Operating temperatures of the STANDARD G6PD Test by site and specimen (degrees Celsius).

| | Overall | HC | MCK |
|---|---|---|---|
| **Venous** | | | |
| Mean temperature (SD) | 26.91 (0.95) | 26.78 (0.95) | 27.04 (0.94) |
| Median temperature | 26.79 | 26.62 | 27.02 |
| Range | 24.11-30.14 | 24.47-29.81 | 24.11-30.14 |
| **Capillary** | | | |
| Mean temperature (SD) | 26.75 (3.34) | 27.05 (3.3) | 26.45 (3.40) |
| Median temperature | 27.46 | 28.17 | 26.67 |
| Range | 17.13-38.87 | 19.41–38.87 | 17.13–38.67 |

**Supplementary Table 2.** Sensitivity and specificity: 2 x 2 table of the STANDARD G6PD Test against the reference G6PD assay for G6PD deficient venous specimens.

| | True positive | True negative | Total |
|---|---|---|---|
| **STANDARD G6PD Test deficient** | 26 | 20 | 46 |
| **STANDARD G6PD Test normal or intermediate** | 0 | 813 | 833 |
| **Total** | 26 | 833 | 859 |

**Supplementary Table 3.** Sensitivity and specificity: 2 x 2 table of the STANDARD G6PD Test against the G6PD reference assay for G6PD intermediate (> 30%–70%), female venous specimens.

| | True positive | True negative | Total |
|---|---|---|---|
| **STANDARD G6PD Test intermediate** | 11 | 18 | 29 |
| **STANDARD G6PD Test normal** | 6 | 286 | 292 |
| **Total** | 17 | 304 | 321 |

**Supplementary Table 4.** Sensitivity and specificity: 2 x 2 table of the STANDARD G6PD Test against the G6PD reference assay for G6PD deficient capillary specimens.

| | True positive | True negative | Total |
|---|---|---|---|
| **STANDARD G6PD Test deficient** | 27 | 21 | 48 |
| **STANDARD G6PD Test normal or intermediate** | 0 | 840 | 840 |
| **Total** | 27 | 861 | 888 |

**Supplementary Table 5.** Sensitivity and specificity: 2 x 2 table of the STANDARD G6PD Test against the G6PD reference assay for G6PD intermediate (> 30%–70%), female capillary specimens.

| | True positive | True negative | Total |
|---|---|---|---|
| **STANDARD G6PD Test intermediate** | 10 | 12 | 22 |
| **STANDARD G6PD Test normal** | 6 | 298 | 304 |
| **Total** | 16 | 310 | 326 |

**Supplementary Table 6.** Sensitivity and specificity: 2 x 2 table of the STANDARD G6PD Test against the G6PD reference assay for G6PD deficient venous specimens (malaria positives only).

| | True positive | True negative | Total |
|---|---|---|---|
| **STANDARD G6PD Test deficient** | 3 | 0 | 3 |
| **STANDARD G6PD Test normal or intermediate** | 4 | 214 | 218 |
| **Total** | 7 | 214 | 221 |

**Supplementary Table 7.** Sensitivity and specificity: 2 x 2 table of the STANDARD G6PD Test against the G6PD reference assay for G6PD intermediate (> 30%–70%), female venous specimens (malaria positives only).

| | True positive | True negative | Total |
|---|---|---|---|
| **STANDARD G6PD Test intermediate** | 1 | 0 | 1 |
| **STANDARD G6PD Test normal** | 3 | 39 | 42 |
| **Total** | 4 | 39 | 43 |

**Supplementary Table 8.** Sensitivity and specificity: 2 x 2 table of the STANDARD G6PD Test against the G6PD reference assay for G6PD deficient capillary specimens (malaria positives only).

| | True positive | True negative | Total |
|---|---|---|---|
| **STANDARD G6PD Test deficient** | 4 | 0 | 4 |
| **STANDARD G6PD Test normal or intermediate** | 7 | 221 | 228 |
| **Total** | 11 | 221 | 232 |

**Supplementary Table 9.** Sensitivity and specificity: 2 x 2 table of the STANDARD G6PD Test against the G6PD reference assay for G6PD intermediate (> 30%–70%), female capillary specimens (malaria positives only).

| | True positive | True negative | Total |
|---|---|---|---|
| **STANDARD G6PD Test intermediate** | 1 | 0 | 1 |
| **STANDARD G6PD Test normal** | 2 | 39 | 41 |
| **Total** | 3 | 39 | 42 |

**Supplementary Table 10.** G6PD status classification: agreement between the STANDARD G6PD Test and the G6PD reference assay using venous specimens.

| | Pointe Scientific reference assay | Total |
|---|---|---|

| | | Deficient | Intermediate | Normal | |
|---|---|---|---|---|---|
| **STANDARD G6PD Test (Venous specimens)** | **Deficient** | 26 | 6 | 14 | 46 |
| | **Intermediate** | 0 | 5 | 15 | 20 |
| | **Normal** | 0 | 6 | 787 | 793 |
| **Total** | | **26** | **17** | **816** | **859** |

Percent agreement: 95.2% (95% CI: 93.6% - 96.6%)

**Supplementary Table 11**. G6PD status classification: agreement between the STANDARD G6PD Test and the G6PD reference assay using capillary specimens.

| | | **Pointe Scientific reference assay** | | | **Total** |
|---|---|---|---|---|---|
| | | **Deficient** | **Intermediate** | **Normal** | |
| **STANDARD G6PD Test (Capillary specimens)** | **Deficient** | 27 | 3 | 18 | 48 |
| | **Intermediate** | 0 | 7 | 7 | 14 |
| | **Normal** | 0 | 6 | 820 | 826 |
| **Total** | | **27** | **16** | **845** | **888** |

Percent agreement: 96.2% (95% CI: 94.7% - 97.3%)

**Supplementary Table 12.** Mean difference and standard deviation by specimen type, for the STANDARD G6PD Test and the HemoCue hemoglobin measurements, compared to the reference complete blood count (CBC) assay.

| **Specimen type** | **Comparison** | **Mean difference** | **Standard deviation** |
|---|---|---|---|
| Capillary | STANDARD G6PD vs CBC | -0.85 | 1.10 |
| | HemoCue vs CBC | -0.10 | 1.04 |
| Venous | STANDARD G6PD vs CBC | 0.43 | 1.21 |
| | HemoCue vs CBC | 0.19 | 0.70 |

**Supplementary Table 13.** Anemia status classification: agreement between the HemoCue and reference CBC's hemoglobin concentration (in g/dL) -based anemia classifications as per WHO definitions on (A) capillary and (B) venous specimens.

**A. Capillary specimens**

| | | **CBC (venous)** | | | |
|---|---|---|---|---|---|
| | | No/mild anemia | Moderate anemia | Severe anemia | Total |
| **HemoCue (capillary)** | No/mild anemia | 326 | 12 | 0 | 338 |
| | Moderate anemia | 27 | 71 | 2 | 100 |
| | Severe anemia | 1 | 5 | 28 | 34 |
| | Total | 354 | 88 | 30 | 472 |

Percent agreement: 90.0% (95% CI: 87.0 – 92.6)

**B. Venous specimens**

| | | **CBC (venous)** | | | |
|---|---|---|---|---|---|
| | | No/mild anemia | Moderate anemia | Severe anemia | Total |
| **HemoCue (venous)** | No/mild anemia | 357 | 8 | 0 | 365 |
| | Moderate anemia | 1 | 81 | 3 | 85 |
| | Severe anemia | 2 | 1 | 27 | 30 |
| | Total | 360 | 90 | 30 | 480 |

Percent agreement: 96.9% (95% CI: 94.9 – 98.2)

**Supplementary Table 14.** Diagnostic performance of the STANDARD G6PD Test and the HemoCue comparator for the identification of severe anemia on (A) capillary and (B) venous blood against the reference complete blood count (CBC) assay on venous blood.

**A. Capillary specimens**

| *Capillary specimens* | | CBC | | |
|---|---|---|---|---|
| | | Severe anemia | Moderate, mild, and no anemia | Total |
| **STANDARD G6PD** | Severe anemia | 25 | 24 | 49 |
| | Moderate, mild, and no anemia | 0 | 417 | 417 |
| | Total | 25 | 441 | 466 |
| Overall agreement: 94.9% (95% CI: 92.4 – 96.7) | | | | |
| | **Sensitivity** | **Specificity** | **PPV** | **NPV** |
| | 100.0% (86.3% – 100.0%) | 94.6% (92.0% – 96.5%) | 51.0% (36.3% – 65.6%) | 100.0% (99.1% – 100.0%) |

| *Capillary specimens* | | CBC | | |
|---|---|---|---|---|
| | | Severe anemia | Moderate, mild ,and no anemia | Total |
| **HemoCue** | Severe anemia | 28 | 6 | 34 |
| | Moderate, mild and no anemia | 2 | 436 | 438 |
| | Total | 30 | 442 | 472 |
| Overall agreement: 98.3% (95% CI: 96.7 – 99.3) | | | | |
| | **Sensitivity** | **Specificity** | **PPV** | **NPV** |
| | 93.3% (77.9% – 99.2%) | 98.6% (97.1% – 99.5%) | 82.4% (65.5% – 93.2%) | 99.5% (98.4% – 99.9%) |

**B. Venous specimens**

| *Venous specimens* | | CBC | | |
|---|---|---|---|---|
| | | Severe anemia | Moderate, mild, and no anemia | Total |
| **STANDARD G6PD** | Severe anemia | 25 | 6 | 31 |
| | Moderate, mild, and no anemia | 1 | 423 | 424 |
| | Total | 26 | 429 | 455 |
| Overall agreement: 98.5% (95% CI: 96.9 – 99.4) | | | | |
| | **Sensitivity** | **Specificity** | **PPV** | **NPV** |
| | 96.2% (80.4% – 99.9%) | 98.6% (97.0% – 99.5%) | 80.6% (62.5% – 92.5%) | 99.8% (98.7% – 100.0%) |

| *Venous specimens* | | CBC | | |
|---|---|---|---|---|
| | | Severe anemia | Moderate, mild ,and no anemia | Total |
| **HemoCue** | Severe anemia | 27 | 3 | 30 |
| | Moderate, mild and no anemia | 3 | 447 | 450 |
| | Total | 30 | 450 | 480 |
| Overall agreement: 98.8% (95% CI: 97.3 – 99.5) | | | | |
| | **Sensitivity** | **Specificity** | **PPV** | **NPV** |
| | 90.0% (73.5% – 97.9%) | 99.3% (98.1% – 99.9%) | 90.0% (73.5% – 97.9%) | 99.3% (98.1% – 99.9%) |

**Supplementary Table 15.** Anemia status* of the study population, by malaria status.

| | Malaria positive | Malaria negative | Total |
|---|---|---|---|
| **No/mild anemia** | 227 (79.6%) | 530 (78.1%) | 757 (79.6%) |
| **Moderate anemia** | 41 (15.1%) | 112 (16.5%) | 153 (16.1%) |
| **Severe anemia** | 4 (1.5%) | 37 (5.5%) | 41 (4.3%) |
| **Total** | 272 (100%) | 679 (100%) | 951 (100%) |

* Anemia status was determined by the HemoCue on venous specimens, applying WHO definitions.

**Supplementary Table 16.** Anemia status* of the study population, by G6PD status.

| | G6PD deficient (<30%) males and females | G6PD intermediate (30-70%) females | G6PD normal males (≥30%) and females (>70%) | Total |
|---|---|---|---|---|
| **No/mild anemia** | 24 (88.9%) | 8 (47.1%) | 725 (79.9%) | 757 (79.6%) |
| **Moderate anemia** | 2 (7.4%) | 9 (52.9%) | 142 (15.7%) | 153 (16.1%) |
| **Severe anemia** | 1 (3.7%) | 0 (0.0%) | 40 (4.4%) | 41 (4.3%) |
| **Total** | 27 (100%) | 17 (100%) | 907 (100%) | 951 (100%) |

*Anemia status was determined by the HemoCue on venous specimens, applying WHO definitions.

**Supplementary Table 17.** Primer sequence for primers used to amplify G6PD exons

| Exon amplified | Mutation | Primer name | Sequence | Expected amplified fragment size (bp) |
|---|---|---|---|---|
| **Exon 6** | Mediterranean | M_F | 5' ACTCCCCGAAGAGGGGTTCAAGG 3' | 542 |
| | | M_R | 5' CCAGCCTCCCAGGAGAGAGGAAG 3' | |
| **Exon 3** | Orissa | O_F | 5' CAGCCACTTCTAACCACACACCT 3' | 308 |
| | | O_R | 5' CCGAAGTTGGCCATGCTGGG 3' | |
| **Exon 9** | Kerala-Kalyan | K_F | 5'CAAGGAGCCCATTCTCT 3' | 253 |
| | | K_R | 5'TGCCTTGCTGGGCCTCG 3' | |
| **Exon 6** | Mahidol | Ma_F | 5' ACTCCCCGAAGAGGGGTTCAAGG 3' | 542 |
| | | Ma_R | 5' CCAGCCTCCCAGGAGAGAGGAAG 3' | |

**Supplementary Table 18.** Study participants with confirmed G6PD mutations as determined by genotyping

| Participant ID | Sex | Zygosity | Mutation | % G6PD activity on reference assay | Venous SDB G6PD activity U/gHb | Capillary SDB G6PD activity U/gHb |
|---|---|---|---|---|---|---|
| HC-0030 | Male | Hemizygous | Orissa | 13.4 | 1.7 | 2.0 |
| HC-0155 | Male | Hemizygous | Orissa | 7.3 | 2.1 | 1.8 |
| HC-0170 | Male | Hemizygous | Orissa | 14.9 | 1.9 | 1.8 |
| HC-0216 | Male | Hemizygous | Mahidol | 13.9 | 2.0 | 1.7 |
| HC-0256 | Female | Heterozygous | Orissa | 49.3 | 8.2 | 7.0 |
| HC-0286 | Male | Hemizygous | Orissa | 15.1 | 2.7 | 2.3 |
| HC-0306 | Male | Hemizygous | Orissa | 10.7 | 2.2 | 2.3 |
| HC-0326 | Male | Hemizygous | Mediterranean | 4.2 | 0.9 | 1.8 |
| HC-0397 | Female | Heterozygous | Orissa | 148.7 | 4.4 | 11.5 |
| HC-0407 | Male | Hemizygous | Orissa | 97.4 | 3.7 | 7.9 |
| HC-0474 | Female | Heterozygous | Orissa | 130.8 | 4.5 | 11.2 |
| HC-0499 | Male | Hemizygous | Mediterranean | 5.2 | 1.4 | 1.2 |
| MCK-0111 | Male | Hemizygous | Orissa | 51.1 | 10 | 13.3 |
| MCK-0123 | Female | Heterozygous | Orissa | 86.2 | 1.7 | 5.7 |
| MCK-0158 | Male | Hemizygous | Orissa | 98.7 | 3.4 | 7.9 |
| MCK-0177 | Female | Homozygous | Orissa | 28.0 | 2.8 | 3.0 |
| MCK-0187 | Male | Hemizygous | Orissa | 6.5 | 1.6 | 1.7 |
| MCK-0357 | Male | Hemizygous | Orissa | 8.3 | 1.5 | 1.4 |

**A. Males- capillary** **B. Males- venous**

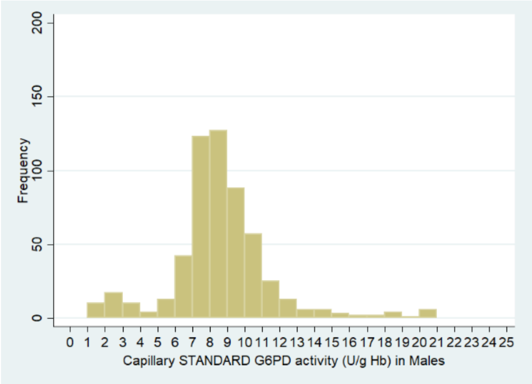


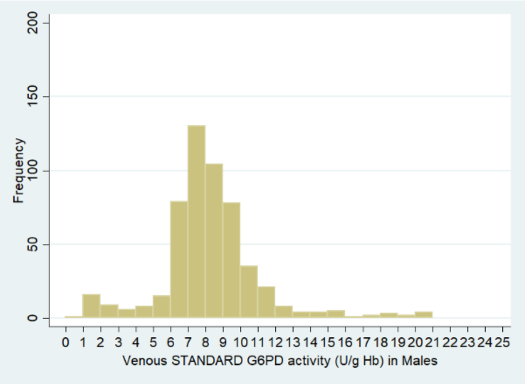


**C. Females- capillary** **D. Females- venous**

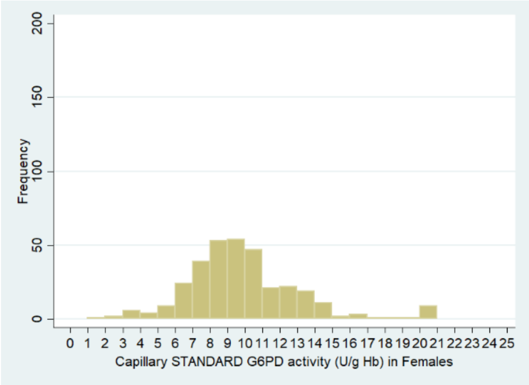


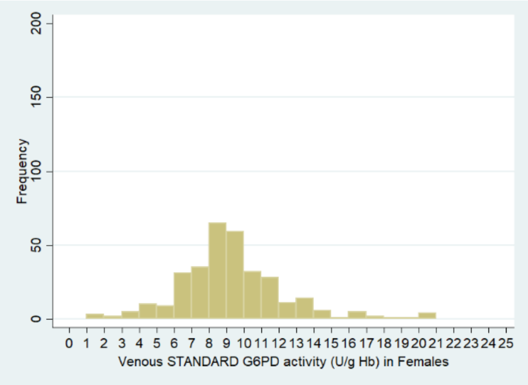


**Supplementary Figure 1.** G6PD activity distributions of study participants on the STANDARD G6PD Test for (A) males- capillary, (B) males- venous, (C) females- capillary, and (D) females- venous.

| A. Deficient males and females, G6PD ≤30% | B. Intermediate females G6PD >30-70% |
|---|---|

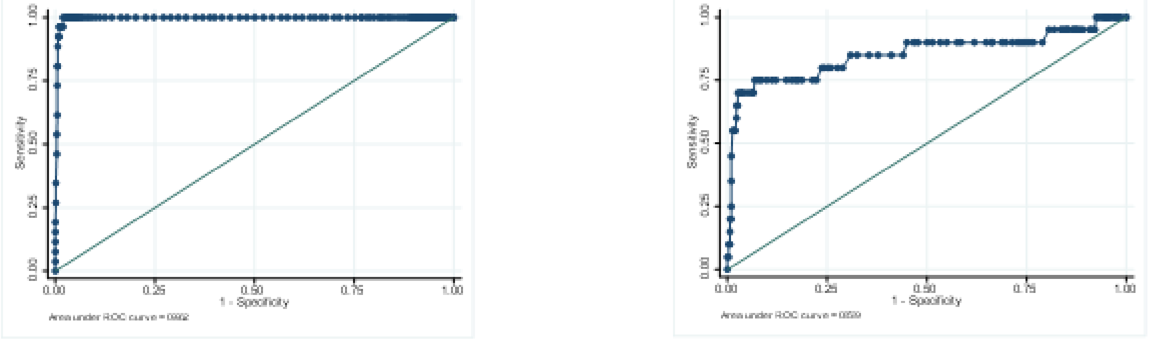


**Supplementary Figure 2.** ROC curves for the STANDARD G6PD measurement on venous specimens and the test's ability to differentiate (A) G6PD normal and deficient males, females from female intermediate; (B) female intermediate with G6PD > 30%–70% from normal females.

**A. Capillary specimens** **B. Venous specimens**

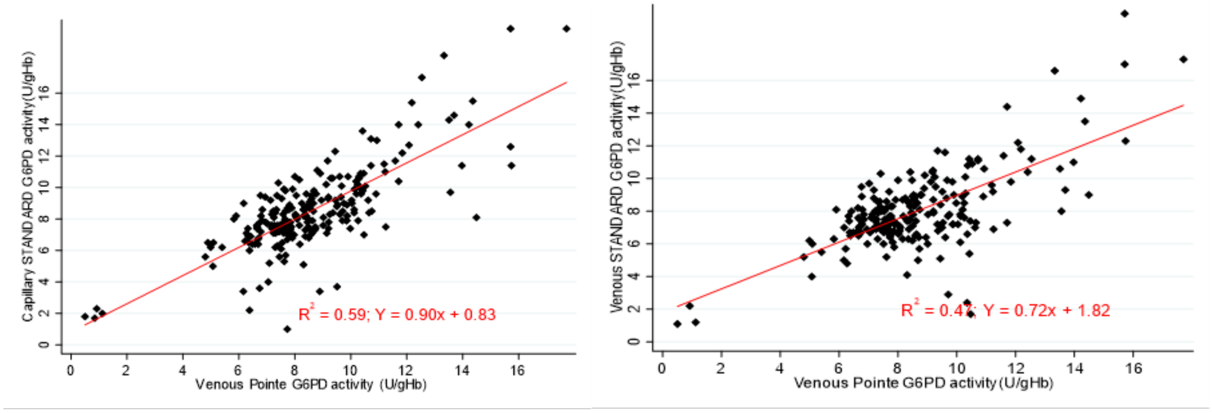


**Supplementary Figure 3.** Correlation of STANDARD G6PD Test by linear regression analysis: G6PD activity on capillary (A) and venous blood (B) versus the Pointe Scientific reference assay on venous blood for malaria positive cases.

### A. Capillary specimens

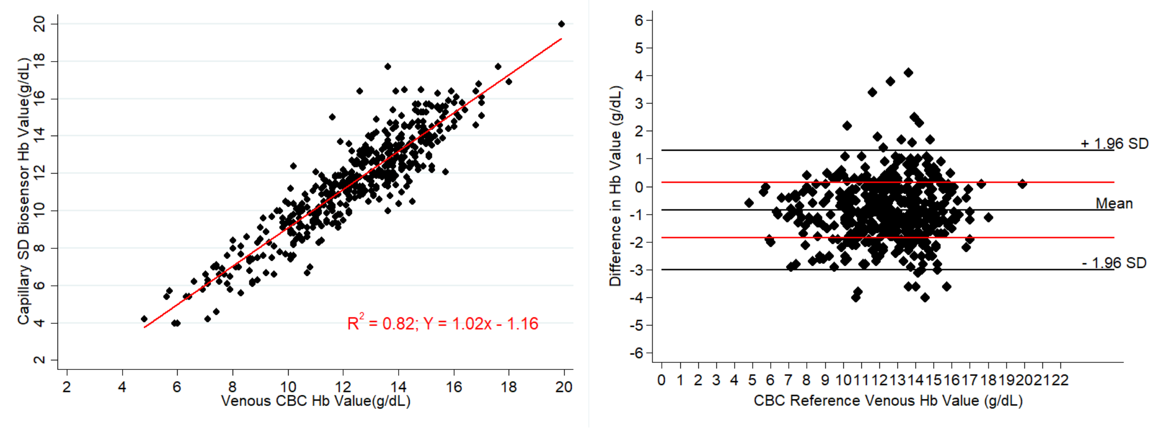


### B. Venous specimens

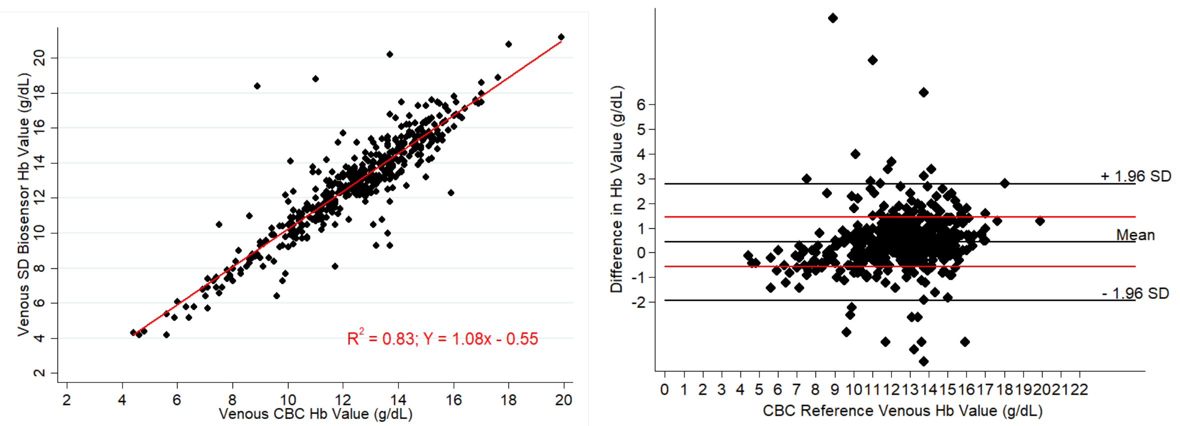


**Supplementary Figure 4.** Correlation of STANDARD G6PD Test hemoglobin results by linear regression analysis and Bland Altmans on capillary (A) and venous blood (B) against the reference complete blood count (CBC) on venous specimens as the reference assay.

**A. Capillary specimens**

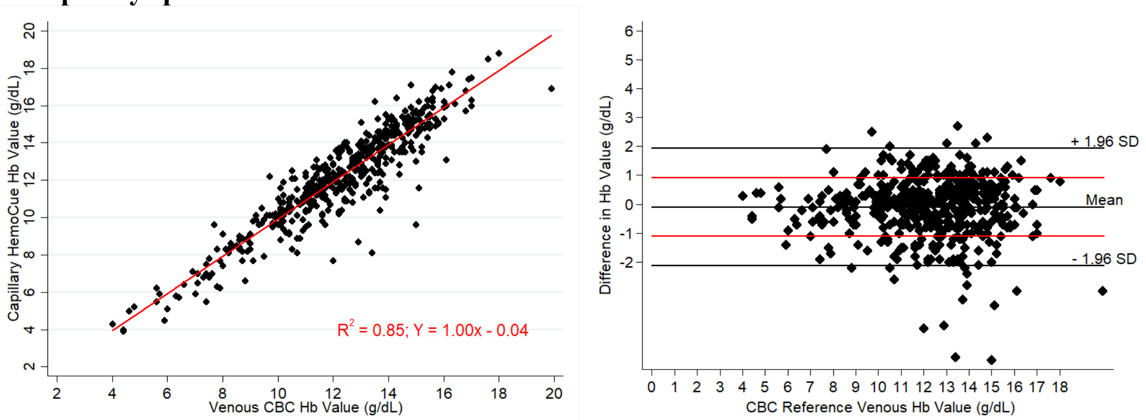


**B. Venous specimens**

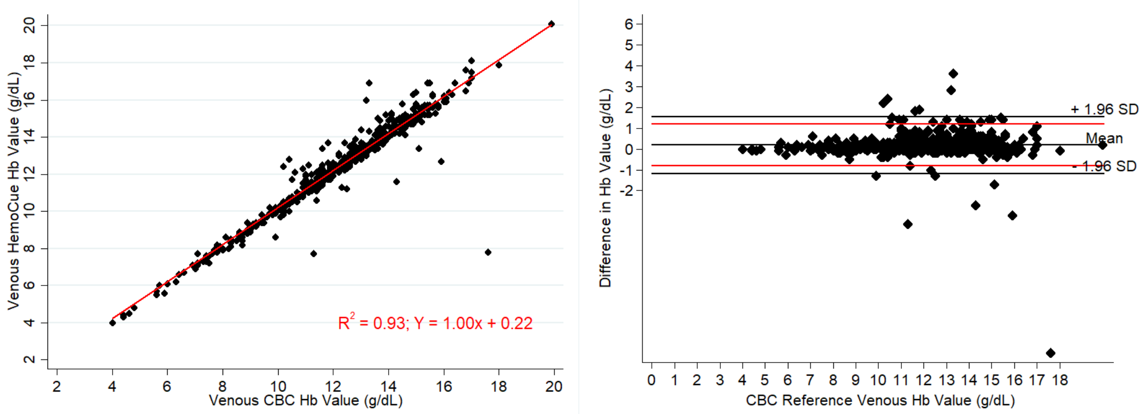


**Supplementary Figure 5.** Correlation of the HemoCue hemoglobin results by linear regression analysis and Bland Altmans on capillary (A) and venous blood (B) against the reference complete blood count (CBC) on venous specimens as the reference assay.
